# Effectiveness of one dose of MVA-BN smallpox vaccine against monkeypox in England using the case-coverage method

**DOI:** 10.1101/2022.12.13.22282654

**Authors:** Marta Bertran, Nick Andrews, Chloe Davison, Bennet Dugbazah, Jacob Boateng, Rachel Lunt, Jo Hardstaff, Melanie Green, Paula Blomquist, Charlie Turner, Hamish Mohammed, Rebecca Cordery, Sema Mandal, Colin Campbell, Shamez N Ladhani, Mary Ramsay, Gayatri Amirthalingam, Jamie Lopez Bernal

**Affiliations:** Immunisations and Vaccine Preventable Diseases Division, UK Health Security Agency; Monkeypox Data, Epidemiology and Analytics Cell, UK Health Security Agency, United Kingdom; Division of Blood Safety, Hepatitis, STIs and HIV, UK Health Security Agency, United Kingdom; Institute for Global Health, University College London, United Kingdom; Paediatric Infectious Diseases Research Group, St. George’s University of London, Cranmer Terrace, London SW17 0RE, United Kingdom

## Abstract

**Background:** Like many other countries worldwide, the UK experienced a national outbreak of monkeypox disease in May 2022, with case numbers rising rapidly, mainly among gay, bisexual and other men who have sex with men (GBMSM). To control the outbreak, Modified Vaccinia Ankara–Bavaria Nordic (MVA-BN), an attenuated smallpox vaccine, was offered to high-risk GBMSM. We assessed the effectiveness of a single MVA-BN dose against monkeypox disease in high-risk GBMSM.

**Methods:** Monkeypox cases in England were sent questionnaires collecting information on demographics, vaccination history and symptoms. Returned questionnaires with a rash onset date (or alternative proxy) between July 04 and October 09, 2022 were included. Females, heterosexual men, and those with missing vaccination information were excluded. Vaccine effectiveness was calculated using the case-coverage method where vaccine coverage among cases is compared to coverage in the eligible population, estimated from doses given to GBMSM and the estimated size of high-risk GBMSM. Sensitivity analysis included +/-20% differences in estimated high-risk GBMSM population size.

**Findings:** Vaccine uptake among eligible GBMSM increased steadily from July 2022, reaching 47% by October 09, 2022. Of the 363 confirmed cases, 8 occurred ≥14 days after vaccination, 32 within 0-13 days after vaccination, and the rest were unvaccinated. The estimated vaccine effectiveness ≥14 days after a single dose was 78% (95% CI: 54%-89%), with a range of ±7% in sensitivity analyses. Vaccine effectiveness within 0–13-days after vaccination was -4% (95% CI: -50% to 29%).

**Interpretation:** A single MVA-BN dose was highly protective against monkeypox disease among high-risk GBMSM.

**Funding:** None

**Research in context:** *Evidence before this study:* We searched PubMed using the terms ‘monkeypox’, ‘MVA’ and ‘vaccine’, with no time limit, and used the snowball process to identify additional relevant publications. We also searched websites of regulatory authorities (FDA, EMA) for any data used during the regulatory approval processes. We also scoped pre-print databases vaccine effectiveness studies during the current outbreak. Only publications related to the Modified Vaccinia Ankara – Bavaria Nordic (MVA-BN) vaccine were included. In the UK, MVA-BN was offered to high-risk GBMSM to control a national outbreak which began in May 2022. MVA-BN is now licensed against smallpox in the US, Europe and the UK, there are, however, limited data on vaccine effectiveness against monkeypox. Preclinical studies indicated two vaccine doses were immunogenic and generated antibody levels considered protective against smallpox. Vaccine-induced antibodies are also cross-protective against monkeypox virus in vitro and in animal models. A recent, as yet unpublished, Israeli study estimated 79% vaccine effectiveness after one dose in high-risk GBMSM, while a US study reported unvaccinated individuals to be 14 times more likely to develop monkeypox disease than vaccinated persons.

*Added value of this study:* Few countries have recommended or introduced large-scale vaccination against the current global outbreak of monkeypox disease among GBMSM in non-endemic countries. The offer of MVA-BN to high-risk GBMSM through sexual health clinics in England provided a unique opportunity to rapidly assess vaccine effectiveness after a single dose using the case-coverage method, which involves comparing vaccine coverage in cases to vaccine coverage in the eligible population. Our vaccine effectiveness estimate of 78% at least 14 days after one MVA-BN dose is consistent with Israeli estimates and provided additional evidence of a lack of protection during the first 13 days after vaccination.

*Implications of all the available evidence:* A single dose of MVA-BN is highly protective against monkeypox disease and provides a useful tool for outbreak control when rapid protection may be needed. Given the lack of effectiveness in the first 13 days after the first dose and a median incubation period of 8-9 days after exposure to the virus, vaccination is likely to be most effective when offered as pre-exposure rather than prophylaxis. Because of the high vaccine effectiveness after one MVA-BN dose, in outbreaks where number of at-risk individuals exceed vaccine supply of two-doses, there may be benefit in prioritising delivery of first doses at the expense of delaying the second dose.

## Introduction

In May 2022, a monkeypox outbreak was identified in the United Kingdom (UK), primarily among gay, bisexual and other men who have sex with men (GBMSM). Similar outbreaks were subsequently identified across Europe and globally,^1,2^ leading the World Health Organisation to declare the outbreak a Public Health Emergency of International Concern (PHEIC) on July 23, 2022.^3^ As of October 11, 2022, more than 20,000 cases have been confirmed in EU/EEA countries (including 4 deaths) as part of this outbreak,^4^ including 3,500 cases in the UK.^5^ Vaccination, with a third generation smallpox vaccine has been a critical component of the outbreak control, yet prior to this outbreak, there has been no data on the clinical effectiveness of the vaccine against monkeypox in humans.

Following the first confirmed cases in England, the UK Health Security Agency (UKHSA) implemented extensive public health measures to control the outbreak, including isolation of cases and close contacts, surveillance of contacts and raising awareness among healthcare professionals and high-risk groups).^6^ Additionally, UKHSA recommended post-exposure vaccination for close contacts of cases, including healthcare workers and laboratory staff, as well as pre-exposure vaccination for healthcare staff who were likely to come into contact with patients with monkeypox disease or monkeypox virus samples.^7^ From June 2022, because of increasing case numbers, the Joint Committee on Vaccination (JCVI) and UKHSA also recommended pre-exposure vaccination for GBMSM considered at higher risk of monkeypox disease during this outbreak,^8^ such as those with multiple partners, participating in group sex and attending sex on premises venues. Proxy indicators, including recent bacterial sexually transmitted infection (in the past year) or eligibility for HIV pre-exposure prophylaxis, were used to identify eligible groups ^7^ with roll-out starting at the end of June 2022 for this group. ^8^

Modified Vaccinia Ankara – Bavaria Nordic (MVA-BN), is a third-generation attenuated replication deficient smallpox vaccine licensed by the European Medicines Agency (EMA) in 2013 for the prevention of smallpox. ^9^ In the US, the Food and Drug Administration (FDA) approved MVA-BN for the prevention of monkeypox as well as smallpox in 2019.^10^ The vaccine was licensed based on immunogenicity studies only; a two dose schedule given 28 days apart was demonstrated to be immunogenic, generating antibody levels above protective thresholds for smallpox, which was also considered to also confer protection against monkeypox. ^11,12^ Animal models have also shown rapid protection against monkeypox. ^13-16^

Although data from studies in Africa suggested that previous generation smallpox vaccination also protected against monkeypox, ^16,17^ there were no real-world data on the effectiveness of third-generation smallpox vaccines against monkeypox disease in humans prior to the current outbreak.^16^ A recent study involving subjects vaccinated during the current outbreak in the Netherlands found relatively low levels of neutralizing antibody titres against monkeypox virus after one dose with no increase in titres after two doses. ^18^ Early reports from Israel and the United States, however, indicate high protection from vaccination against monkeypox disease.^19 20^

Following implementation of a large-scale national immunisation programme for at-risk GBMSM, we undertook an observational study to estimate the effectiveness of one dose of MVA-BN against symptomatic monkeypox disease in England.

## METHODS

We estimated vaccine effectiveness in the GBMSM cohort eligible for MVA-BN vaccination using the case-coverage method (also known as the screening method), whereby vaccination rates among cases is compared to population coverage. ^21^

### Population vaccine coverage

Weekly vaccinations delivered to the -at risk GBMSM population along with an estimate of the GBMSM denominator was used to estimate coverage. The number of GBMSM at high risk of monkeypox has previously been estimated as 111,000 in the UK. We used the same calculation to estimate an England total of 89,240. This is based on an estimated 48,500 GBMSM regular sexual health clinic attenders at high risk of monkeypox and inflation factors of an additional 60% (29,100) non regular attenders and on top of this another 15% (11,640) not in contact with sexual health services. ^22^ To account for uncertainty this denominator was increased and decreased by 20% in sensitivity analyses.

An aggregated vaccination coverage reporting system was established at a vaccination site level at the start of the programme rollout by NHS England.^23^ Vaccination sites, primarily sexual health clinics, reported daily number of vaccines delivered by cohort (i.e. GBMSM, healthcare workers, contacts of cases) and by dose (supplementary figure S1). We classified vaccine status as recent (vaccinated in the current or previous week) or one full dose (vaccinated with a first dose at least 2 weeks earlier). At the time of analysis, very few second doses had been delivered so second dose vaccination was not assessed.

### Vaccination rates among cases

Vaccination status of cases was obtained from questionnaires sent to confirmed (Monkeypox positive) and highly probable (Orthopox positive) cases via email and/or text message,^24^ which also included questions on symptoms, rash onset, age, gender and sexual orientation. When available, personal identifiers such as name and date of birth were also used to link cases to other electronic systems (HP zone – a public health case management system, laboratory results) to obtain missing and additional information, such as symptom onset and laboratory specimen date (supplementary figure S1).

For each case an index date for monkeypox infection was defined in the following priority order depending on whether the previous information was available

a. Date of rash onset
b. Date of symptom onset from the questionnaire
c. Date of symptom onset from HP zone
d. Date of test from the lab minus 4 days
e. Date of test from the questionnaire minus 4 days
f. Date of questionnaire completion minus 14 days

Dates d to e were based on median interval seen in those with a date of rash and date of test or questionnaire.

Only cases with an index date from July 04, 2022, when routine GBMSM scaled up (figure 1) until the week commencing 3 October 2022 were included.

**Figure 1.**
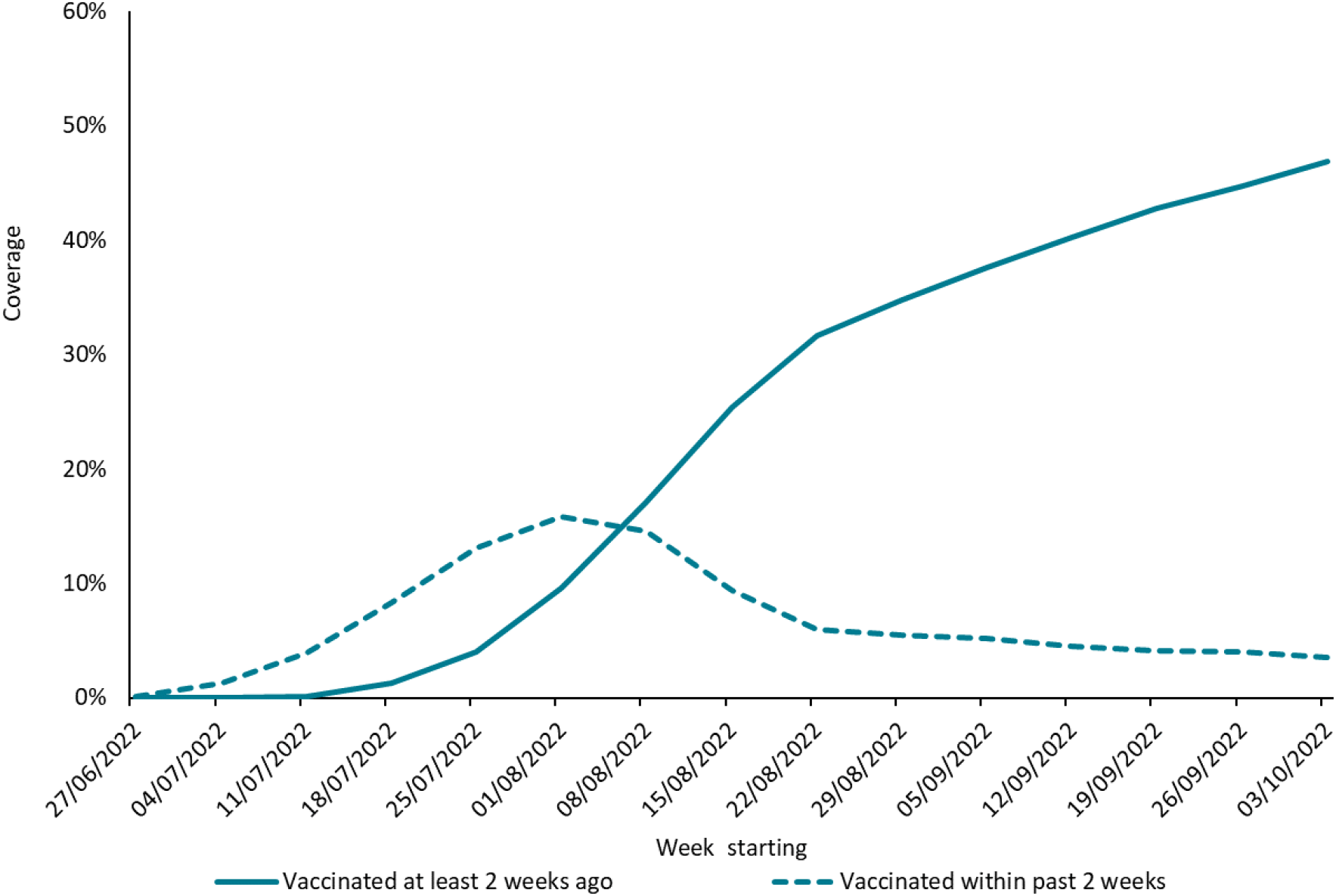
Estimated first dose coverage by week for recent MVA-BN vaccinations and vaccinations at least 2 weeks ago in gay, bisexual and other men who have sex with men at high risk of monkeypox in England, weeks 26-40

Vaccination status was categorised as being either pre-1971, 1971-2022, in 2022 (with dates given) or not known/missing. Cases vaccinated after the index date were included as unvaccinated. Cases reporting vaccination in 2022 but without a date were only included in a sensitivity analysis because they may have been vaccinated after disease. Cases vaccinated prior to 2022 were considered unvaccinated – genuine vaccinations prior to 2022 were most likely to include childhood smallpox vaccinations, and the UK ended smallpox vaccination in 1971.

To estimate the proportion of cases with completed questionnaires, we used a numerator of returned questionnaires where the diagnostic sample date was between weeks 28-38 (July 11 - September 25). For the denominator we used the number of cases sent a text message with a link to the questionnaire during this period (figure 1). Note that week 27 cases by sample date were mainly not eligible for the study (as they likely had rash onset prior to week 27) and cases in weeks 39 and 40 had not had sufficient time for all questionnaires to be returned. An exact return rate could not be calculated as it was not always possible to link questionnaires sent and returned because some questionnaires were completed anonymously.

### Statistical method

Vaccine effectiveness (VE) is calculated as 1-odds of vaccination in cases/ odds of vaccination in the population. ^21,25^ This was done using the weekly aggregated cases matched to the appropriate vaccine coverage and analysed using a logistic regression model with an offset for the log-odds of the coverage and a fitted constant. VE is 1-the exponent of the estimated parameter.^25^ When calculating VE in those at least 14 days earlier, those vaccinated within 0-13 days are not included. Consequently, vaccine coverage for this cohort was calculated as the proportion of the cohort vaccinated at least 14 days previously)/(1-proportion vaccinated within 0-13 days). A similar adjustment was made when calculating corrected coverage for those vaccinated within 0-13 days (i.e., subtracting the ≥14 days coverage). Analyses were performed using Stata v17.0 (StatCorp, Tx).

To account for the impact that prior smallpox vaccination might have in individuals vaccinated against smallpox during childhood, VE was also calculated for cases under the age of 50. However, we used the overall population coverage in this analysis as these data are not available by age.

### Ethical approval

The study was conducted in accordance with the relevant guidelines and regulations under permissions granted to UK Health Security Agency (UKHSA, formerly Public Health England) under Regulation 3 of The Health Service (Control of Patient Information) Regulations 2020 and under Section 251 of the NHS Act 2006 (United Kingdom legislation). Specifically, all data were collected within the statutory approvals granted to the UKHSA for infectious disease surveillance and control. Information was held securely and in accordance with the Data Protection Act 2018 (United Kingdom legislation) and Caldicott Principles (UK, https://www.gov.uk/government/publications/the-caldicott-principles).

## RESULTS

### Population coverage

Vaccine coverage by week for first doses increased to 47% by early October (week 40), with most vaccinations being given by August 22, 2022 (week 34). Analysis of protection within 2 weeks of vaccination included the cohort that was mostly vaccinated during July and August 2022(Figure 1). The number of doses given and estimated coverage by week (recent and ≥14 days since) are included in supplementary table S2. Cases were matched to coverage based on their index week, for example a case with index week 33 matches to recent vaccination coverage of 9.4% and vaccination at least 2 weeks ago of 25.4%. For the sensitivity analysis reducing the denominator by 20% increases coverage at the end of September to 63% and increasing it by 20% reduces coverage to 42%.

### Cases

A total of 1,102 cases had responded to questionnaires by November 03, 2022; 52 were female or self-declared male heterosexuals and excluded from the analysis. Only one of these 52 had reported to have been vaccinated with an unknown date in 2022 and this case’s onset was in week 26 prior to our study period. Overall, 460 cases had an index date from July 04, 2022, which was based on rash onset date for 412, other symptoms for 32, test-4 days for 4 and questionnaire completion date -14 days for 12 cases. Their vaccination status is summarised in Table 1.

**Table 1.** Vaccination status of cases by age group and classifications for analysis

| **Year of vaccination** | **Interval of vaccination before index date** | **In analysis as** | **<50 years** | **≥50 years** | **Unknown age** | **Total** |
| --- | --- | --- | --- | --- | --- | --- |
| **Not vaccinated** | Not applicable | Unvaccinated in 2022 | 217 | 38 | 260 | 260 |
| **Before 1971** | Many years | Unvaccinated in 2022 | 3 | 30 | 0 | 29 |
| **1971 to 2021** | Years | Unvaccinated in 2022 | 24 | 4 | 1 | 33 |
| **2022** | 0-13 days before rash | Recent vaccination | 26 | 3 | 0 | 29 |
| **2022** | 0-13 before non-rash index date | Recent vaccination | 3 | 0 | 0 | 3 |
| **2022** | ≥14 days before rash | 1 dose | 4 | 0 | 0 | 4 |
| **2022** | ≥14 days before non-rash index date | 1 dose | 3 | 1 | 0 | 4 |
| **2022** | Vaccine date missing | Potential 1 dose | 1 | 0 | 0 | 1 |
| **Not known/prefer not to say** | Not applicable | Dropped | 67 | 25 | 5 | 97 |
| **Total** |  |  | **348** | **101** | **11** | **460** |

After excluding cases with an unknown vaccination date (n=97), the total included in the analyses was 363 cases. Four cases were vaccinated ≥ 14 days before rash onset and a further four cases were likely to have been vaccinated ≥14 days prior to infection. For these four cases the index date was based on symptom onset date for two and test-4 days for the other two. The interval between vaccination and monkeypox disease were longer than 3 weeks for these 4 cases. The one case with the missing vaccination date missing was included in the sensitivity analysis (Table 1). No cases reported receiving two vaccine doses. Figure 2 shows the number of cases by week according to vaccination status within those who returned a questionnaire and gave information on vaccination status. Lower numbers in the latest weeks reflect declining incidence and lower questionnaire return rate.

**Figure 2.**
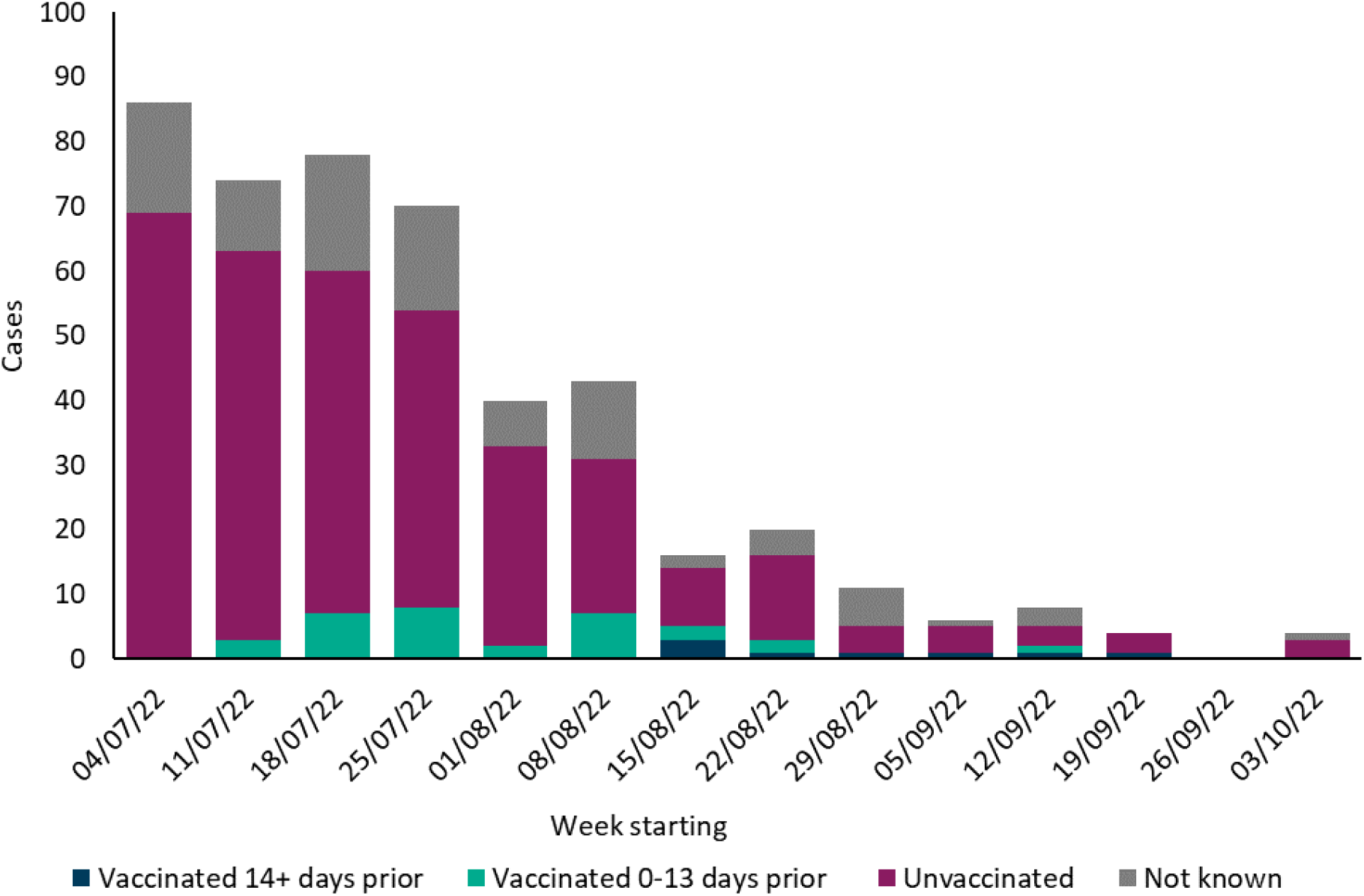
Number of monkeypox cases by week and vaccination status from July 04, 2022 (week 27) to October 09, 2022 (week 40)

The age of the 460 cases returning the questionnaire and their vaccine status is shown in table 2. Individuals over 50 years old are potentially eligible for smallpox vaccination as an infant. This age group accounts for 21% of cases (table 2), compared to 14% of all England cases in men (data not shown). As expected, proportionally more of those over 50 reported vaccination before 2022 (table 1). There was only one vaccine failure in this age.

**Table 2.** Age distribution of cases by vaccine status

| Age | Vaccine status unknown | Vaccine status known | Total |
| --- | --- | --- | --- |
| 15-19 | 1 | 4 | 5 |
| 20-24 | 6 | 18 | 24 |
| 25-29 | 10 | 45 | 55 |
| 30-34 | 11 | 60 | 71 |
| 35-39 | 9 | 56 | 65 |
| 40-44 | 18 | 59 | 77 |
| 45-49 | 12 | 39 | 51 |
| 50-54 | 17 | 29 | 46 |
| 55-59 | 6 | 19 | 25 |
| 60+ | 1 | 28 | 30 |
| Unknown | 5 | 6 | 11 |
| **Total** | **97** | **363** | **460** |

### Estimated questionnaire return rate

Between weeks 28-38 (July 11-September 25) a total of 2018 cases were reported in England of whom an 1545 (77%) had questionnaires sent by text. A total of 508 questionnaires were returned in this period giving a return rate of 33%.

### 3.2 Vaccine effectiveness

The central estimate of VE after a single dose of smallpox vaccine was 78% (95% CI: 54%-89%) (Table 3). In the sensitivity analyses, reducing the GBMSM denominator by 20% resulted in higher population vaccine coverage and, therefore, a higher VE of 85% (95% CI: 69% to 93%), while increasing the GBMSM denominator by 20% reduced VE to 71% (95% CI: 40% to 86%) (Table 3). When the extra case with unknown vaccine date was added to those vaccinated ≥14 days prior, VE dropped to 75% (95% CI: 50%-87%). Single-dose VE within 0-13 days was -4% (95% CI: -50% to 29%) for the central estimate and ranged from -30% (95% CI: -89% to 10%) to 23% (95% CI: -11% to 47%), not different to zero in any scenario (Table 3). In the analysis including only cases aged under 50, VE was 74% (95% CI: 43%-88%) (table 3).

**Table 3.** Vaccine effectiveness (VE) estimates using the screening method for all ages and for cases under 50 years of age, including results from sensitivity analyses using different denominators

| Age group | Total number of gay, bisexual and other men who have sex with men assumed to be at high risk of monkeypox | Dose 1 Interval | Cases (n/%) | Matched Coverage* | VE (95% CI) |
| --- | --- | --- | --- | --- | --- |
| All ages | 89,240 | 0 to 13 days | 32 (8.8%) | 7.9% | -4% (-50% to 29%) |
|  |  | ≥14 days | 8 (2.2%) | 8.0% | 78% (54% to 89%) |
|  |  | unvaccinated | 322 (89.0%) | 84.1% | - |
| All ages | 71,392 (-20%) | 0 to 13 days | 32 (8.8%) | 9.9% | 23% (-11% to 47%) |
|  |  | ≥14 days | 8 (2.2%) | 10.0% | 85% (69% to 93%) |
|  |  | unvaccinated | 322 (89.0%) | 80.1% | - |
| All ages | 107,088 (+20%) | 0 to 13 days | 32 (8.8%) | 6.6% | -30% (-89% to 10%) |
|  |  | ≥14 days | 8 (2.2%) | 6.7% | 71% (40% to 86%) |
|  |  | unvaccinated | 322 (89.0%) | 86.7% | - |
| <50 | 89,240 | 0 to 13 days | 29 (10.4%) | 8.1% | -21% (-80% to 18%) |
|  |  | ≥14 days | 7 (2.5%) | 7.8% | 74% (43% to 88%) |
|  |  | unvaccinated | 244 (87.1%) | 84.1% | - |
| <50 | 71,392 (-20%) | 0 to 13 days | 29 (10.4%) | 10.1% | 10% (-33% to 33%) |
|  |  | ≥14 days | 7 (2.5%) | 9.8% | 82% (61% to 92%) |
|  |  | unvaccinated | 244 (87.1%) | 80.1% | - |
| <50 | 107,088 (+20%) | 0 to 13 days | 29 (10.4%) | 6.7% | -52% (-126% to -3%) |
|  |  | ≥14 days | 7 (2.5%) | 6.5% | 66% (25% to 84%) |
|  |  | unvaccinated | 244 (87.1%) | 86.8% | - |

\*Coverage matched by index date week, this is a lot below final cover as many cases occurred in the early weeks of roll out. VE: Vaccine effectiveness; 95%CI: 95% confidence intervals

## Discussion

We estimated MVA-BN vaccine effectiveness against monkeypox disease at least 14 days after a single dose to be 78% (range 71%-85% in sensitivity analyses), with no evidence of protection in the first 13 days after vaccination.

Our estimates are consistent with the recent Israeli study, which estimated a VE of 79% (95% CI: 24%-94%) after a single dose of MVA-BN in a similar cohort of high-risk GBMSM aged 18-42 years using an electronic healthcare system that covers around 52% of the Israeli population.^19^ The eligible cohort in this study was similar to the cohort in our study: male subjects at high risk of infection, defined by similar criteria to those in England.^19^ Only subjects aged 18-42 were including, likely excluding any subjects who might have received childhood smallpox vaccinations, which would be comparable to our <50 analysis. The analysis did not report VE within and after 14 days, with a follow-up interval of 25 days after vaccination. Similarly, a US study reported a 14-fold higher incidence of monkeypox disease in unvaccinated individuals eligible for MVA-BN compared to those receiving at least vaccine dose in 32 jurisdictions, covering 56% of the US population eligible for vaccination, with similar trends observed in a sensitivity analysis but lower relative incidence rates. ^20^ This risk is equivalent to a vaccine effectiveness of around 93%. Similarly to our study, the authors used aggregate data provided by public health jurisdictions, which obtained vaccination date from case interviews and/or linkage to vaccine registries. ^26^ They restricted their analysis by age to exclude anyone who might have been vaccinated with the smallpox vaccination in childhood (those aged 50+ years), as we did in our secondary analysis. We also focused on VE at least 14 days after vaccination to allow time for an immune response to develop after vaccination. The higher VE in the US study may in part be explained by the fact that they excluded those with possible prior smallpox vaccination (which we cannot exclude from our population comparator), and they also included a small number who had received second doses. Another study conducted in France, focused on subjects who received post-exposure vaccination, researchers reported 12/276 (4%) had a confirmed monkeypox breakthrough infection.^26^ Of these 12 cases, 10 were within 1-5 days of vaccination. This highlights the importance of considering the period soon after vaccination separately in our study.

Our effectiveness results after one dose are also consistent with pre-clinical studies based on immunogenicity responses and animal models.^12,14^ One of these studies assessed the response after one single dose of MVA in animal models challenged with monkeypox.^14^ Full protection was shown after 30 days but antibody response and protection against severe disease (and death) was shown as soon as 4 days after vaccination – although the sample size was small.^14^ During this outbreak, Zaeck et al showed low levels of MPXV neutralizing antibodies after one dose of vaccine in a small sample of human subjects. ^18^ Interestingly, there was a difference between individuals born before or after 1974 – with 63% of the younger group eliciting neutralizing antibodies compared with 100% of those born before 1974, ^18^ who were likely to have received previous smallpox vaccination – showing additional protection against monkeypox. These individuals also showed higher levels of neutralizing antibodies than those born after 1974 - unlikely to have received any previous smallpox vaccination. ^18^ Correlates of protection for monkeypox are unclear and further studies are required including serology and clinical outcomes. However, this study suggests the need to differentiate between subjects previously vaccinated during childhood and unvaccinated individuals. This is in line with reports from surveillance data from Africa suggesting a protective effect of previous smallpox vaccination, in some cases estimated to be about 85% effective. ^16,17^ In line with these results, our analysis in cases under 50 years of age, which are less likely to have been vaccinated prior to this outbreak, also showed slightly lower VE.

There are some limitations to this analysis. Firstly, we assumed all cases are vaccine eligible GBMSM unless they reported as female or heterosexual. A small number of cases may not have been eligible for vaccination. Given the high percentage of all cases in England self-reporting as GBMSM (96.8%)^22^, however, this should not have a measurable influence in the final analysis. We relied on questionnaire data to obtain case vaccine status. The questionnaire return rate was low (about 33%) and, potentially, if vaccinated cases are more likely to return completed questionnaires, VE would be underestimated and *vice versa*. Vaccine information was obtained only from self-reported data, there is no national vaccination registry with identifiable data for this programme. There is also some uncertainty on the GBMSM denominator, resulting in a range of VE from 75%-87% in the sensitivity analyses. Given the aggregated nature of the vaccine coverage data, we were also unable to adjust for other potential confounders other than time period. This could lead to VE being overestimated or underestimated. For example, if cases within the GBMSM population included in the study were younger, but vaccine uptake was higher in older individuals then VE would be overestimated (and note that the age of cases included in the study compared to all cases was slightly younger, but we do not know if coverage differed by age). Behavioural changes post-vaccination may also have affected VE estimates; for example, if vaccinated individuals were less likely to abstain from high-risk sexual activity as a result of having received the vaccine then the pharmacological effectiveness would be underestimated. We considered individuals vaccinated prior to 2022 as unvaccinated because we could only assess the effect of vaccination as part of the 2022 immunisation campaign since it was not possible to split vaccine coverage by prior vaccine status. If prior vaccination has some residual protection, then this means that our estimates may be lower than the true VE. The fact that VE was similar when restricting to cases aged under 50 who were less likely to have received a vaccine prior to 2022 suggests the bias may be fairly small. However, many cases are in people born outside of the UK, including from regions where smallpox vaccination programmes may have ended later than 1971. Also, some GBMSM cases might be healthcare workers or laboratory workers who might have received vaccination. Therefore, this age cut-off still includes some cases who had been previously vaccinated. In fact, 7% of <50s reported vaccination between 1971 and 2021 but this figure likely reflects recall bias given the very low eligibility since 1971.^27^

It is important to note that most of these potential biases would also affect our VE estimates during 0-13 day after vaccination. The fact this estimate is close to zero provides some reassurance that these potential biases are unlikely to be having a major impact on our VE estimates for ≥14 days post-vaccination.

Finally, vaccination using an intradermal vaccination route instead of subcutaneous to maximise the number of doses was recommended in the UK from August 22, 2022, with variable rollout dates across the country and with variable implementation between clinics. ^7^ By November 2022, 65% of sites were offering intradermal vaccinations. Consequently, most doses in our analysis would have been administered subcutaneously and, therefore, our VE estimates relate mainly to subcutaneous vaccinations. Any VE differences between subcutaneous and intradermal administration will need to be assessed in future studies.

## Conclusions

Our results suggest relatively high level of protection from a single dose of MVA-BN – making it a useful tool against monkeypox for outbreak control when rapid protection is needed. This suggests that where numbers at highest risk of infection exceed vaccine supply of two-doses, there may be benefit in prioritising delivery of first doses. Further work is needed to evaluate the duration of protection as well as the effectiveness of a full two dose course.

## Data Availability

All data produced in the present work are contained in the manuscript.

## Acknowledgements

We would like to thank NHS England for their collection and sharing of vaccination coverage rates for the monkeypox vaccination programme delivered through sexual health services. We would also like to thank UK Health Security Agency Health Protection Teams for sending out the questionnaires as part of the public health management and surveillance of cases and the cases themselves for completing these questionnaires.

---

**Figure S1:**
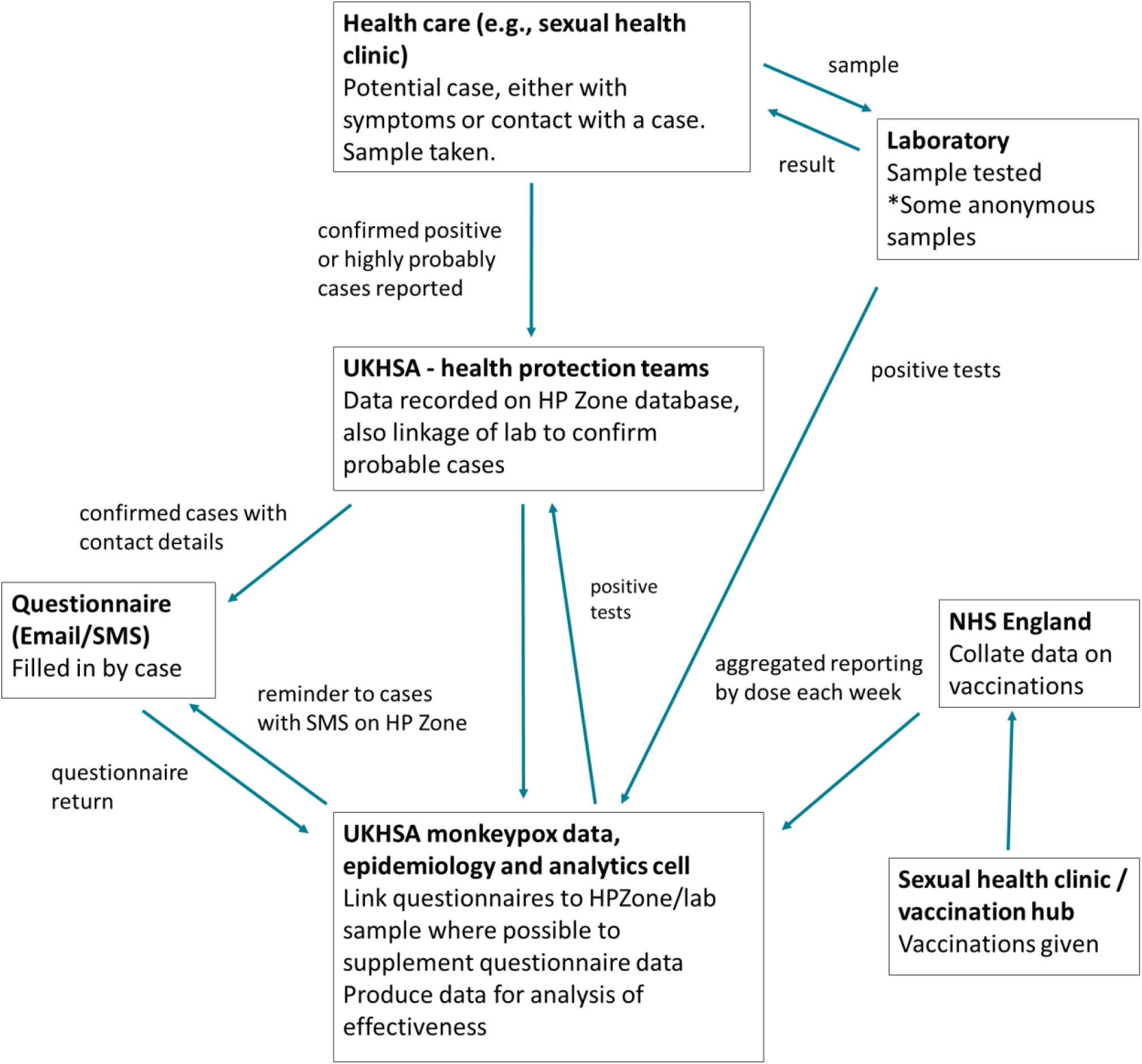
Flowchart

## References

1. Vaughan AM, Cenciarelli O, Colombe S, Alves de Sousa L, Fischer N, Gossner CM, et al. A large multi-country outbreak of monkeypox across 41 countries in the WHO European Region, 7 March to 23 August 2022. Euro Surveill. 2022;27(36).

2. Thornhill JP, Barkati S, Walmsley S, Rockstroh J, Antinori A, Harrison LB, et al. Monkeypox Virus Infection in Humans across 16 Countries - April-June 2022. N Engl J Med. 2022;387(8):679–91.

3. World Health Organization. News: WHO Director-General declares the ongoing monkeypox outbreak a Public Health Emergency of International Concern. 2022 [Available from: https://www.who.int/europe/news/item/23-07-2022-who-director-general-declares-the-ongoing-monkeypox-outbreak-a-public-health-event-of-international-concern.

4. ECDC. Monkeypox situation update, as of 11 October 2022 2022 [Available from: https://www.ecdc.europa.eu/en/news-events/monkeypox-situation-update#:~:text=Since%20the%20start%20of%20the,Western%20Balkan%20countries%20and%20Turkey.

5. UK Health Security Agency. Monkeypox outbreak: epidemiological update, 4 October 2022 2022 [Available from: https://www.gov.uk/government/publications/monkeypox-outbreak-epidemiological-overview/monkeypox-outbreak-epidemiological-overview-4-october-2022.

6. UK Health Security Agency. Monkeypox: guidance. [Available from: https://www.gov.uk/government/collections/monkeypox-guidance.

7. UK Health Security Agency. Recommendations for the use of pre and post-exposure vaccination during a monkeypox incident. 2022 [Available from: https://assets.publishing.service.gov.uk/government/uploads/system/uploads/attachment_data/file/1100600/recommendations-for-pre-and-post-exposure-vaccination-during-a-monkeypox-incident-26-august-2022.pdf.

8. UK Health Security Agency. Monkeypox outbreak: vaccination strategy 2022 [Available from: https://www.gov.uk/guidance/monkeypox-outbreak-vaccination-strategy#pre-exposure-vaccination.

9. European Medicines Agency. Imvanex: smallpox and monkeypox vaccine (Live Modified Vaccinia Virus Ankara). [Available from: https://www.ema.europa.eu/en/medicines/human/EPAR/imvanex.

10. Food and Drug Administration. FDA approves first live, non-replicating vaccine to prevent smallpox and monkeypox. 2019 [Available from: https://www.fda.gov/news-events/press-announcements/fda-approves-first-live-non-replicating-vaccine-prevent-smallpox-and-monkeypox.

11. Frey SE, Newman FK, Kennedy JS, Sobek V, Ennis FA, Hill H, et al. Clinical and immunologic responses to multiple doses of IMVAMUNE (Modified Vaccinia Ankara) followed by Dryvax challenge. Vaccine. 2007;25(51):8562–73.

12. Pittman PR, Hahn M, Lee HS, Koca C, Samy N, Schmidt D, et al. Phase 3 Efficacy Trial of Modified Vaccinia Ankara as a Vaccine against Smallpox. N Engl J Med. 2019;381(20):1897–908.

13. Earl PL, Americo JL, Wyatt LS, Eller LA, Whitbeck JC, Cohen GH, et al. Immunogenicity of a highly attenuated MVA smallpox vaccine and protection against monkeypox. Nature. 2004;428(6979):182–5.

14. Earl PL, Americo JL, Wyatt LS, Espenshade O, Bassler J, Gong K, et al. Rapid protection in a monkeypox model by a single injection of a replication-deficient vaccinia virus. Proc Natl Acad Sci U S A. 2008;105(31):10889–94.

15. Hatch GJ, Graham VA, Bewley KR, Tree JA, Dennis M, Taylor I, et al. Assessment of the protective effect of Imvamune and Acam2000 vaccines against aerosolized monkeypox virus in cynomolgus macaques. J Virol. 2013;87(14):7805–15.

16. Poland GA, Kennedy RB, Tosh PK. Prevention of monkeypox with vaccines: a rapid review. Lancet Infect Dis. 2022.

17. Fine PE, Jezek Z, Grab B, Dixon H. The transmission potential of monkeypox virus in human populations. Int J Epidemiol. 1988;17(3):643–50.

18. Zaeck LM, Lamers MM, Verstrepen BE, Bestebroer TM, van Royen ME, Götz H, et al. Low levels of monkeypox virus neutralizing antibodies after MVA-BN vaccination in healthy individuals. Nature Medicine. 2022.

19. Ronen Arbel YWS, Roy Zucker et al. [Pre-print] Effectiveness of a single-dose Modified Vaccinia Ankara in Human Monkeypox: an observational study. Research Square. 2022.

20. Payne AB RL, Kugeler KJ, et al. Incidence of Monkeypox Among Unvaccinated Persons Compared with Persons Receiving ≥1 JYNNEOS Vaccine Dose — 32 U.S. Jurisdictions, July 31–September 3, 2022. MMWR Morb Mortal Wkly Rep. 2022;71:1278–1282.

21. Farrington CP. Estimation of vaccine effectiveness using the screening method. Int J Epidemiol. 1993;22(4):742–6.

22. UK Health Security Agency. Investigation into monkeypox outbreak in England: technical briefing 8. 2022 [Available from: https://www.gov.uk/government/publications/monkeypox-outbreak-technical-briefings/investigation-into-monkeypox-outbreak-in-england-technical-briefing-8.

23. NHS England. Vaccinations for monkeypox. 2022 [Available from: https://www.england.nhs.uk/statistics/statistical-work-areas/vaccinations-for-monkeypox/.

24. UK Health Security Agency. Monkeypox: Case definitions 2022 [Available from: https://www.gov.uk/guidance/monkeypox-case-definitions.

25. Parikh SR, Andrews NJ, Beebeejaun K, Campbell H, Ribeiro S, Ward C, et al. Effectiveness and impact of a reduced infant schedule of 4CMenB vaccine against group B meningococcal disease in England: a national observational cohort study. Lancet. 2016;388(10061):2775–82.

26. Thy M, Peiffer-Smadja N, Mailhe M, Kramer L, Ferré VM, Houhou-Fidouh N, et al. Breakthrough infections after post-exposure vaccination against Monkeypox. medRxiv. 2022:2022.08.03.22278233.

27. UK Health Security Agency. The Green Book: Chapter 29: Smallpox and monkeypox [Available from: https://assets.publishing.service.gov.uk/government/uploads/system/uploads/attachment_data/file/1106454/Green-Book-chapter-29_Smallpox-and-monkeypox_26September2022.pdf.

