## Supplemental Table S2 for "Effectiveness of one dose of MVA-BN smallpox vaccine against monkeypox in England using the case-coverage method"

**Table S2: Vaccine first doses and coverage estimation by week in GBMSM, England**

| Week number | Week commencing | First doses | Doses 0-13 days ago | Cumulative doses | Doses ≥14 days ago | Denominator | Coverage 0-13 days ago | Coverage ≥14 days ago |
|---|---|---|---|---|---|---|---|---|
| **26** | 27/06/2022 | 90 | 90 | 90 | 0 | 89,240 | 90 | 90 |
| **27** | 04/07/2022 | 1,050 | 1,140 | 1,140 | 0 | 89,240 | 1,050 | 1,140 |
| **28** | 11/07/2022 | 2,439 | 3,489 | 3,579 | 90 | 89,240 | 2,439 | 3,489 |
| **29** | 18/07/2022 | 4,939 | 7,378 | 8,518 | 1,140 | 89,240 | 4,939 | 7,378 |
| **30** | 25/07/2022 | 6,762 | 11,701 | 15,280 | 3,579 | 89,240 | 6,762 | 11,701 |
| **31** | 01/08/2022 | 7,401 | 14,163 | 22,681 | 8,518 | 89,240 | 7,401 | 14,163 |
| **32** | 08/08/2022 | 5,567 | 12,968 | 28,248 | 15,280 | 89,240 | 5,567 | 12,968 |
| **33** | 15/08/2022 | 2,800 | 8,367 | 31,048 | 22,681 | 89,240 | 2,800 | 8,367 |
| **34** | 22/08/2022 | 2,492 | 5,292 | 33,540 | 28,248 | 89,240 | 2,492 | 5,292 |
| **35** | 29/08/2022 | 2,388 | 4,880 | 35,928 | 31,048 | 89,240 | 2,388 | 4,880 |
| **36** | 05/09/2022 | 2,259 | 4,647 | 38,187 | 33,540 | 89,240 | 2,259 | 4,647 |
| **37** | 12/09/2022 | 1,754 | 4,013 | 39,941 | 35,928 | 89,240 | 1,754 | 4,013 |
| **38** | 19/09/2022 | 1,897 | 3,651 | 41,838 | 38,187 | 89,240 | 1,897 | 3,651 |
| **39** | 26/09/2022 | 1,676 | 3,573 | 43,514 | 39,941 | 89,240 | 1,676 | 3,573 |
| **40** | 03/10/2022 | 1,463 | 3,139 | 44,977 | 41,838 | 89,240 | 1,463 | 3,139 |

Pre-print
